# Polygenic associations and causal inferences between serum immunoglobulins and amyotrophic lateral sclerosis

**DOI:** 10.1101/2020.04.07.20057265

**Authors:** Xu Chen, Xiaojun Shen, Xuzhuo Zhang, Yiqiang Zhan, Fang Fang

## Abstract

Chronic inflammation might contribute to the development of amyotrophic lateral sclerosis (ALS), the relationship between serum immunoglobulins and risk of ALS remains however unclear. In order to overcome limitations like reverse causation and residual confounding commonly seen in the observational studies, we applied molecular epidemiological analyses to examine the polygenic and causal associations between serum immunoglobulins and ALS. Summary statistics from the large-scale genome-wide association studies (GWAS) among European ancestry populations (∼15000 individuals for serum immunoglobulins, and more than 36000 individuals for ALS) were accessed from different consortia. The relationships between three types of serum immunoglobulins (IgA, IgM, and IgG) and ALS were investigated in a discovery phase and then in a replication phase. Polygenic risk score (PRS) analysis was performed with PRSice package to test the polygenic association, and Mendelian randomization (MR) analysis was performed with TwoSampleMR package to infer the causality. An inverse polygenic association was discovered between IgA and ALS as well as between IgM and ALS. Such associations were however not replicated using a larger GWAS of ALS, and no causal association was observed for either IgA-ALS or IgM-ALS. A positive polygenic association was both discovered [odds ratio (OR) = 1.18, 95% confidence interval (CI): 1.12-1.25, P=5.9×10^−7^] and replicated (OR=1.13, 95% CI: 1.06-1.20, P=0.001) between IgG and ALS. A causal association between IgG and ALS was also suggested in both the discovery (OR=1.06, 95%CI: 1.02-1.10, P=0.009) and replication (OR=1.07, 95%CI: 0.90-1.24, P=0.420) analyses, although the latter was not statistically significant. This study suggests a shared polygenic risk between serum IgG (as a biomarker for chronic inflammation) and ALS.

## Introduction

Amyotrophic lateral sclerosis (ALS) is a relatively rare but fatal neurodegenerative disease. Chronic inflammation and altered immune responses have been suggested to contribute to the pathogenesis of ALS [1]. The relationship between serum immunoglobulins, as known biomarkers for inflammation and immune responses, and risk of ALS remains however unclear. In a previous study [2], we did not find a statistically significant positive association between immunoglobulin G (IgG) and risk of ALS [hazard ratio=1.04, 95% confidence interval (CI): 0.89-1.22; per 2.99 g/L increase of IgG]. In the same study, we also found that during the 20 years before diagnosis, patients with ALS had more rapidly declining levels of IgG compared to others. We speculate that the noted lack of association between IgG and ALS might be attributable to both a real positive association between IgG and ALS (i.e., higher levels of IgG lead to higher risk of ALS) and reverse causation (i.e., ALS results in declined levels of IgG). In addition to reverse causation, observational studies are also prone to other systemic errors, which collectively make it often difficult to infer causality for noted associations [3].

By using the summary statistics from large-scale genome-wide association studies (GWAS), polygenic risk score (PRS) and Mendelian randomization (MR) analyses have added novel evidence to support polygenic and causal relationships between several traits and ALS, corroborating importantly findings from previous observational studies [4-6]. In this study, we used GWAS summary statistics to assess the associations of genetically predicted levels of immunoglobulins with the genetically predicted risk of ALS, and to test whether such associations are causal. Since IgG is a biomarker for chronic inflammation, we hypothesized that there is a positive and causal relationship between IgG and ALS. We additionally studied another two serum immunoglobulins (IgA and IgM, two biomarkers that have rarely been studied in ALS) to assess the specificity of the IgG result.

## Methods

### Summary statistics

We used publicly available summary statistics of several GWAS within the European ancestry population in this study. GWAS of serum immunoglobulins (IgA, IgM, and IgG) were conducted among ∼15000 individuals (from Iceland and Sweden), from which single nucleotide polymorphisms (SNPs) with the association P-values <1.0×10^−6^ were available [7]. GWAS summary statistics of ALS were obtained from two studies (available in the IEU GWAS database; https://gwas.mrcieu.ac.uk, ID for ALS_2016: ieu-a-1085; ID for ALS_2018: ebi-a-GCST005647). In the discovery analysis, summary statistics of ALS (ALS_2016) were obtained from a GWAS performed among 36052 individuals (including 12577 patients with ALS and 23475 controls) by the Project MinE group [8]. In the replication analysis, summary statistics of ALS (ALS_2018) were obtained from a meta-analysis incorporating the results from ALS_2016 and another GWAS including 40598 individuals (8229 patients with ALS and 32369 controls) [9]. Because we only used GWAS summary statistics rather than individual-level data in this study, participant inform consent and ethical review permit were waived according to the ethical review board of Shenzhen Baoan Women’s and Children’s Hospital.

Using IgA, IgM, and IgG as the exposures (base traits) and ALS as the outcome (target trait), the polygenic and causal associations of six exposure-outcome pairs (three in the discovery analysis and three in the replication analysis) were investigated by PRS and MR analyses, respectively. Because the values of immunoglobulins were standardized in the GWAS, we estimated accordingly odds ratio (OR) per standard deviation of genetically predicted level for each type of immunoglobulin in the present study.

### PRS analyses

Polygenic associations were investigated by PRS analyses in Linux with PRSice package (version 1.25) [10]. For the summary statistics of each exposure, correlated SNPs were pruned by linkage disequilibrium (LD) clumping with default parameter settings. Genotypes of the HapMap_ceu_all (release 22, 60 individuals, 3.96 million SNPs) were used as the reference panel, clumping threshold p1=p2=0.5, LD threshold r^2^=0.05, and the distance threshold of 300Kb. Independent SNPs were then extracted and included in 100000 quantiles with gradually increasing P-value thresholds (P_T_, ranging from 0 to 1.0×10^−6^, in steps of 1.0×10^−11^ per quantile). The P_T_ of the quantile that explained the largest variance of the target was defined as the best-fitted P_T_.

### MR analyses

Utilizing the instrumental variable (IV) methods (Figure 1), MR analyses were performed in R (version 3.6.1) with TwoSampleMR package (version 0.5.2) [11]. Causal inference by MR analysis requires three basic assumptions: 1) relevance, i.e., IVs have causal effects on the exposure (e.g., IgG); 2) exclusion restriction, i.e., IVs affect the outcome (e.g., ALS), only through the exposure; and 3) exchangeability or independence, i.e., no common causes exist between IVs and the outcome [12]. Lead SNPs from the genetic loci reported to be associated with each exposure were extracted from the GWAS of immunoglobulins [7]. The associations of the SNPs with other traits (associations with a P-value <5×10^−8^) were checked in GWAS Catalog (https://www.ebi.ac.uk/gwas) and excluded, leaving independent lead SNPs after LD clumping as the IVs (Table 1). The causal relationship of each exposure-outcome pair was primarily examined by the inverse variance weighted (IVW) method and complemented with another four methods (MR Egger regression, weighted median, simple and weighted mode) as sensitivity analyses.

**Table 1.** SNPs used as instrumental variables for serum immunoglobulins in the MR analyses.

|  | SNP (IV) | Position (hg 38) | Gene | EA | OA | Beta | SE | P-value |
| --- | --- | --- | --- | --- | --- | --- | --- | --- |
| **IgA** | rs35668054 | Chr2:71827707 | *CYP26B1* | T | C | -0.18 | 0.02 | $1.60 \times 10^{-20}$ |
| | rs968567 | Chr11:61828092 | *FADS2* | T | C | 0.12 | 0.01 | $1.30 \times 10^{-17}$ |
| | rs9276244 | Chr6:32733444 | *HLA-DQA2* | G | A | 0.09 | 0.01 | $2.50 \times 10^{-14}$ |
| | rs1071888 | Chr22:29841373 | *ASCC2* | A | T | -0.08 | 0.01 | $3.20 \times 10^{-10}$ |
| | rs10065637 | Chr5:56143024 | *ANKRD55* | T | C | -0.08 | 0.01 | $5.00 \times 10^{-10}$ |
| | rs142973694 | Chr6: 31395164 | *MICA* | A | G | 0.13 | 0.02 | $6.70 \times 10^{-10}$ |
| **IgM** | rs2511713 | Chr8:102565637 | *ODF1* | G | A | -0.13 | 0.01 | $1.70 \times 10^{-24}$ |
| | rs735665 | Chr11:123490689 | *GRAMD1B* | A | G | 0.15 | 0.01 | $1.00 \times 10^{-23}$ |
| | rs6037651 | Chr20:3705789 | *SIGLEC1* | C | T | -0.09 | 0.01 | $2.90 \times 10^{-15}$ |
| | rs617119 | Chr3:32414275 | *CMTM7* | C | T | -0.09 | 0.01 | $1.10 \times 10^{-14}$ |
| | rs11651596 | Chr17:39899863 | *GSDMB* | C | T | -0.09 | 0.01 | $2.30 \times 10^{-14}$ |
| **IgG** | rs7554873 | Chr1:161642443 | *FCGR2B* | C | T | -0.28 | 0.02 | $8.90 \times 10^{-32}$ |
| | rs2133037 | Chr6:32699819 | *HLA-DQB1* | A | G | -0.19 | 0.02 | $2.40 \times 10^{-14}$ |

EA=effect allele; OA=other allele; SE=standard error, which was calculated by using P-value and sample size; P-value=association P-value.

**Figure 1.**
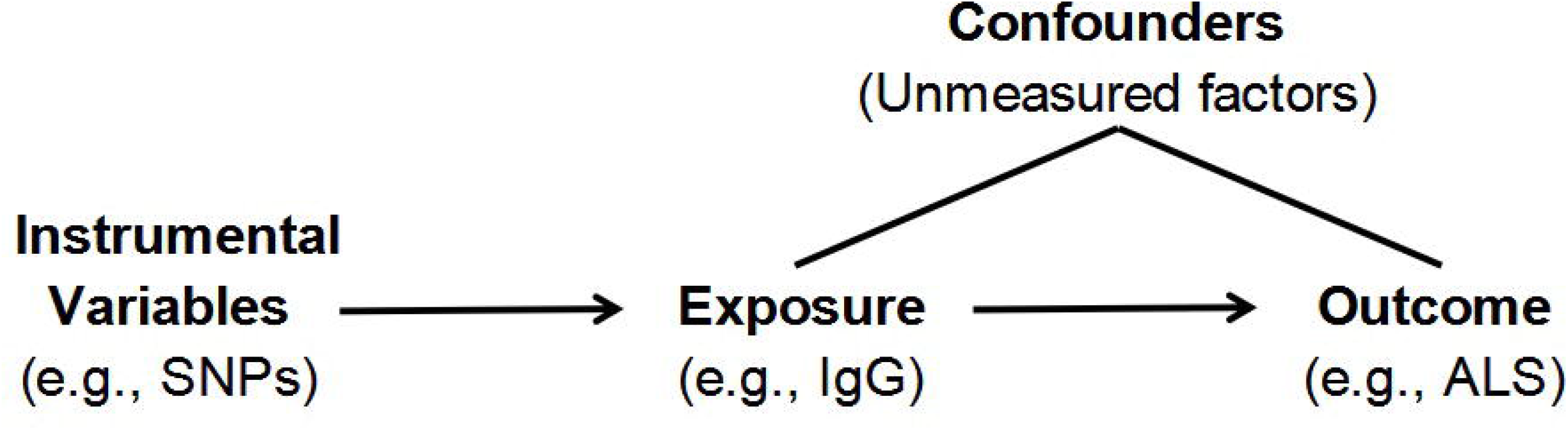
Illustration of the Mendelian randomization analysis. Mendelian randomization analysis utilizes the instrumental variable method to infer the causality between exposure and outcome. Because alleles are randomly assigned during mitosis, restricted single nucleotide polymorphisms (SNPs) could be used as instrumental variables to test the causal effect of exposure on the outcome.

## Results

PRS results between serum immunoglobulins and ALS were presented in Figure 2, which including 10 out of 100000 quantiles in discovery and replication phases, estimates of the best-fitted quantiles were summarized in Table 2. Inverse polygenic associations were discovered for IgA-ALS and IgM-ALS, neither of the associations was however replicated. A positive polygenic association between IgG and ALS was found both in the discovery and replication analyses, the OR was 1.18 (95% CI: 1.12-1.25, P=5.9×10^−7^) in the discovery analysis and 1.13 (95%CI: 1.06-1.20, P=0.001) in the replication analysis.

**Table 2.**
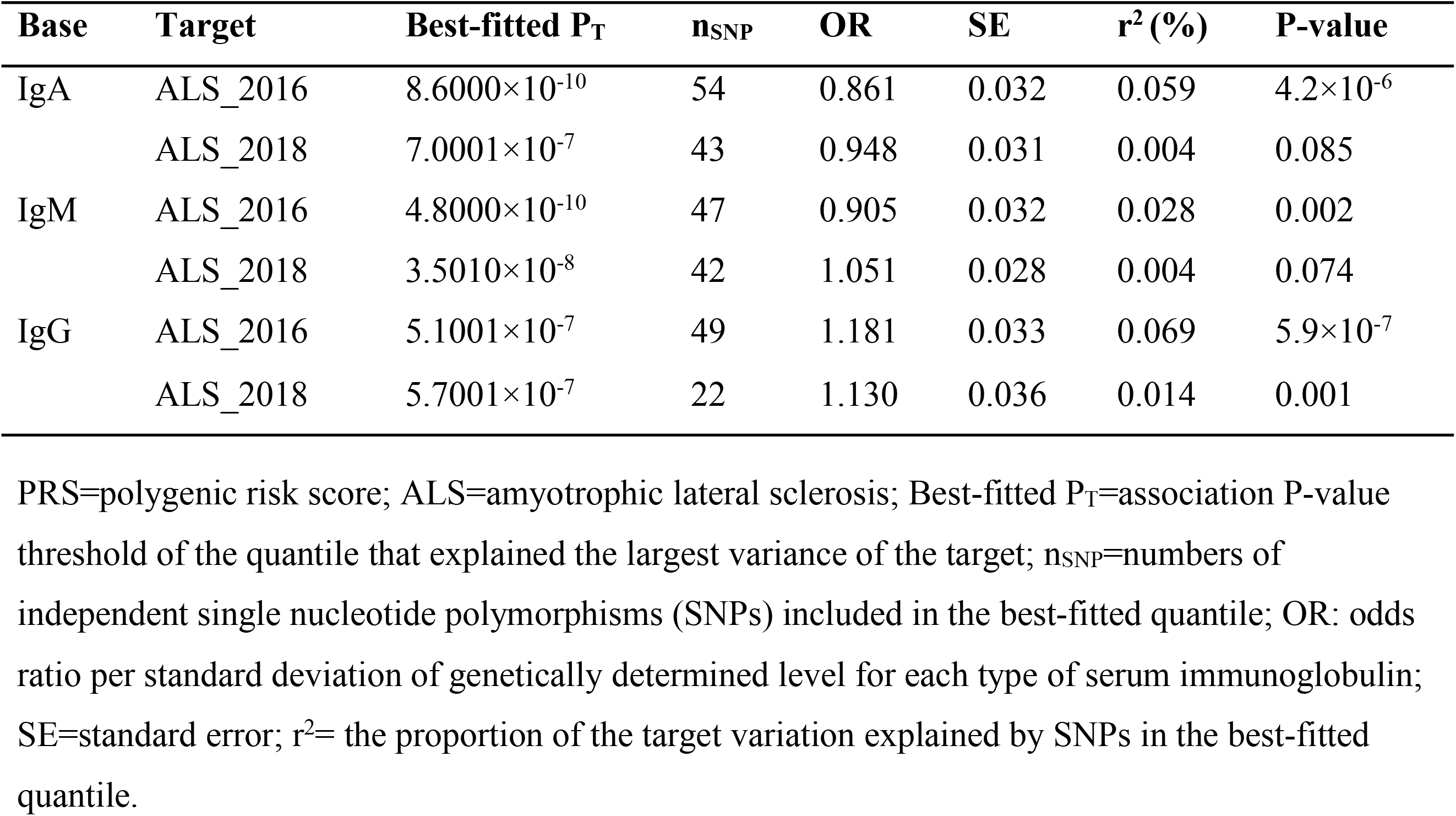
Polygenic associations between serum immunoglobulins and ALS in the PRS analyses.

**Figure 2.**
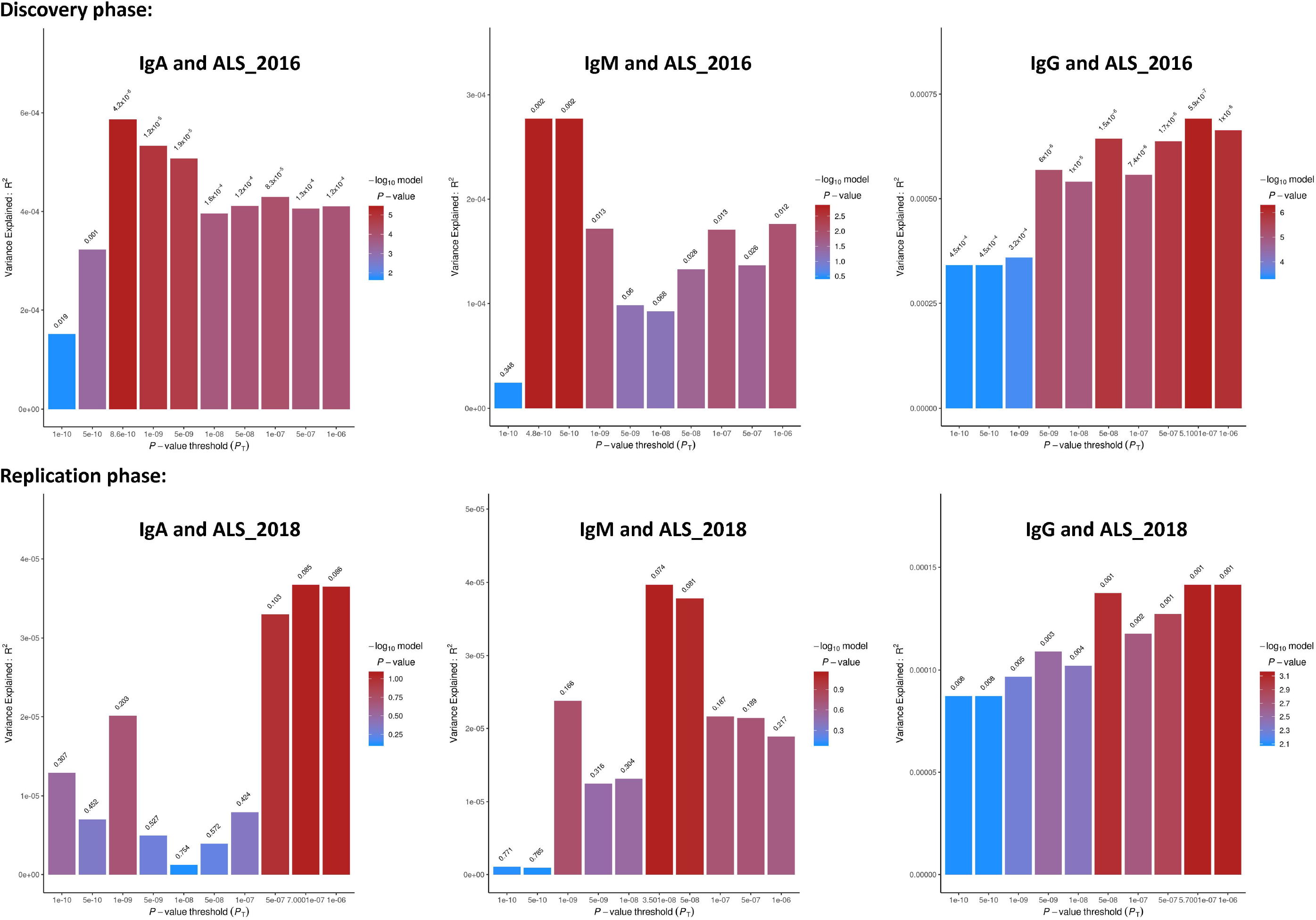
Polygenic risk score analyses between serum immunoglobulins and ALS. Standardized polygenic risk scores based on the alleles increasing each type of immunoglobulin (base trait) are used to predict ALS (target trait). X axis: independent SNPs are extracted from the GWAS of base trait and then included in 100000 quantiles with gradually increasing P-value thresholds (P_T_, ranging from 0 to 1.0×10^−6^, in steps of 1.0×10^−11^ per quantile). The P_T_ of the quantile that explained the largest variance of the target is defined as the best-fitted P_T_. Ten out of 100000 quantiles (including the best-fitted quantile) are presented. Y axis: Nagelkerke r^2^, the proportion of the target variation explained by independent SNPs in the P-threshold quantile.

MR results using different methods to infer the causality between serum immunoglobulins and ALS were presented in Figure 3 and summarized in Table 3, respectively. Among these five MR methods, standard error of estimate from IVW method was the smallest (Table 3), while the MR Egger estimates were less precise (with the largest standard error), because the MR Egger method allows variants to have pleiotropic effects. However, causal association was not found for either IgA-ALS or IgM-ALS, regardless of the use of MR methods. For IgG-ALS, a statistically significant causal association was found in the discovery analysis (OR=1.06, 95% CI: 1.02-1.10, P=0.009). A causal positive association was also suggested in the replication analysis, although not statistically significant (OR=1.07, 95%CI: 0.90-1.24, P=0.420).

**Table 3.** Comparison of results from different methods in Mendelian randomization analyses.

| Exposure | Method | NIV | ALS_2016 |  |  | ALS_2018 |  |  |
| --- | --- | --- | --- | --- | --- | --- | --- | --- |
|  |  |  | OR | SE | Pval | OR | SE | Pval |
| **IgA** | IVW | 6 | 0.968 | 0.022 | 0.148 | 0.880 | 0.100 | 0.201 |
|  | MR Egger | 6 | 1.126 | 0.067 | 0.152 | 1.477 | 0.303 | 0.267 |
|  | Weighted<br>median | 6 | 0.979 | 0.025 | 0.382 | 0.927 | 0.105 | 0.466 |
|  | Simple mode | 6 | 0.990 | 0.035 | 0.783 | 1.033 | 0.180 | 0.866 |
|  | Weighted<br>mode | 6 | 0.991 | 0.030 | 0.774 | 1.008 | 0.152 | 0.960 |
| **IgM** | IVW | 5 | 0.996 | 0.015 | 0.787 | 1.031 | 0.060 | 0.617 |
|  | MR Egger | 5 | 0.962 | 0.070 | 0.619 | 0.873 | 0.279 | 0.661 |
|  | Weighted<br>median | 5 | 0.990 | 0.019 | 0.575 | 1.003 | 0.073 | 0.965 |
|  | Simple mode | 5 | 0.992 | 0.025 | 0.753 | 0.997 | 0.096 | 0.979 |
|  | Weighted<br>mode | 5 | 0.990 | 0.022 | 0.708 | 0.997 | 0.089 | 0.978 |
| **IgG** | IVW | 2 | **1.056** | **0.021** | **0.009** | 1.073 | 0.087 | 0.420 |

NIV=numbers of independent SNPs used as instrumental variables; IVW= inverse variance weighted method; OR: odds ratio per standard deviation of genetically determined level for each type of serum immunoglobulin; SE=standard error; Pval=P-value for the causal inference, significance is defined as Pval<0.05.

**Figure 3.**
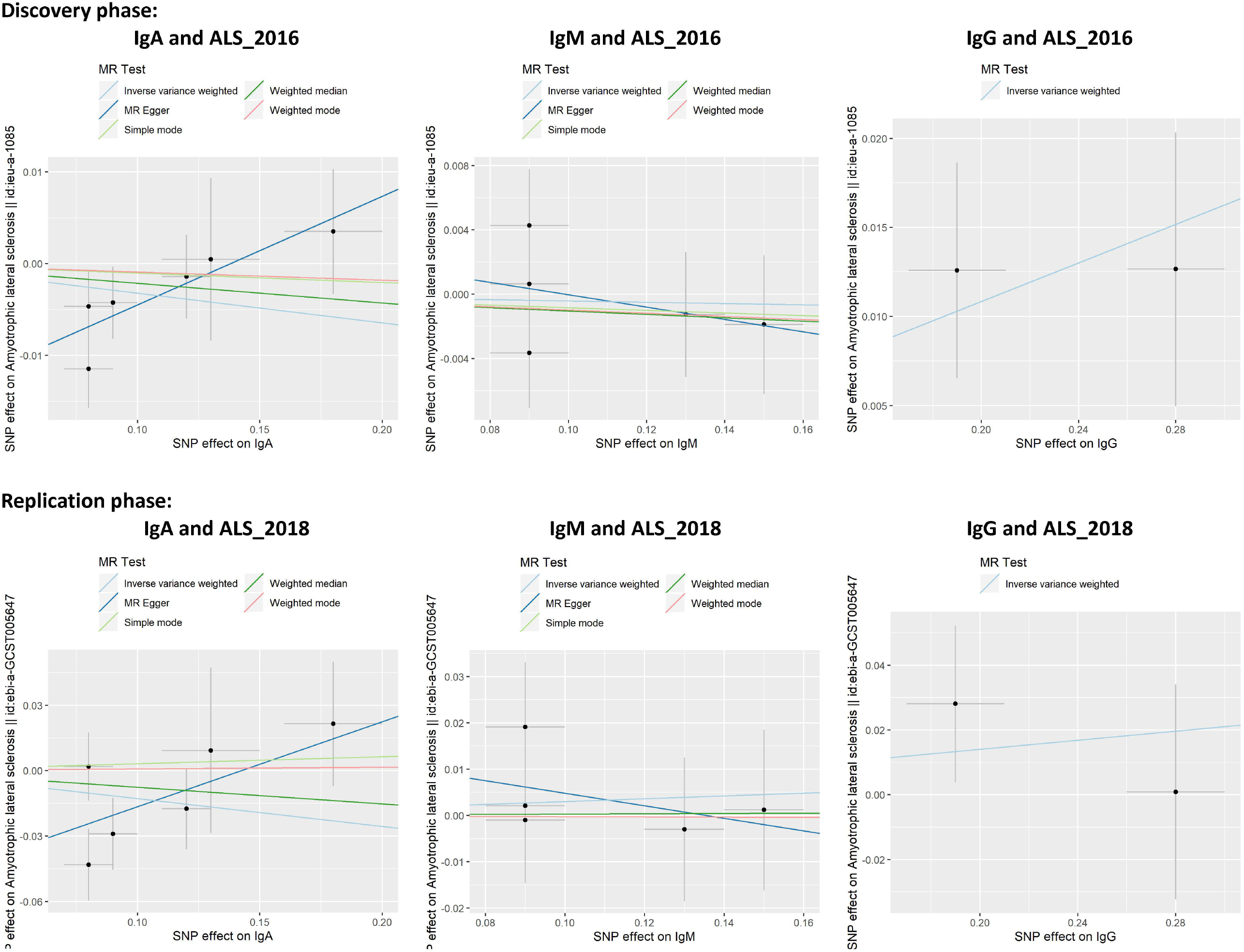
Mendelian randomization analyses between serum immunoglobulins and ALS. Causal inference is performed by Mendelian randomization analysis, using the instrumental variable method. It is primarily examined by the inverse variance weighted (IVW) method and complemented with four other instrumental variable methods (MR Egger regression, weighted median, simple mode, and weighted mode) as sensitivity analyses. X axis: SNP effect on the exposure trait. Y axis: SNP effect on the outcome trait.

## Discussion

Based on summary statistics data from large-scale GWAS of immunoglobulins (IgA, IgM, IgG) and ALS, our PRS analyses provided new evidence supporting a polygenic association between IgG and ALS. The MR analyses suggested potentially causal association between IgG and ALS, but it didn’t reach statistical significance in the replication phase. The inconsistence of MR estimates in discovery and replication phases probably due to weak instrument bias, population stratification or other confounding.

Accumulating evidence supports that inflammation and altered immune responses are involved in the different phases of ALS [13]. Physiological barriers (e.g., blood-brain barrier) and innate immunity cells (e.g., macrophages) are fundamental in the maintenance and modulation of the central nervous system, whereas chronic inflammation might lead to dysfunction of the neurons [14]. B cells play important roles in adaptive immunity, through producing immunoglobulins and presenting antigens to T cells [15]. IgA constitutes 10-20% of serum immunoglobulins and is mainly involved in the mucosal immune response. IgM constitutes 5-10% of serum immunoglobulins and is the earliest released antibody during adaptive immune response. IgG constitutes ∼75% of serum immunoglobulins and can be transmitted through placental and blood-brain barriers [16].

Several studies have indicated a link between IgG and ALS. Deposit of IgG has been found in the spinal cord and motor cortex of some ALS patients [17]. Prevalence of serum monoclonal immunoglobulins (60%, including 44% IgG and 16% IgM) was found to be higher among ALS patients than that of the control group (13%) [18]. High levels of 20 IgG antibodies were also found to help diagnose ALS cases [19]. In our previous cohort study including more than half a million participants with >20 years of follow-up, we found that per 2.99 g/L increase of IgG levels in blood, there was a 4% increased risk of ALS, although the association was not statistically significant [2]. The lack of statistical significance might be multifactorial, including first a potentially real positive association between IgG and ALS risk, a lack of statistical power due to the relatively small number of ALS cases identified (N=152), and also reverse causation (i.e., patients with ALS had more rapidly declining IgG levels, compared to individuals not developing ALS, during the 20 years before ALS diagnosis). However, because genotypes are determined at birth and unchangeable during the life course, our results about a potentially causal relationship between genetically predicted IgG level and genetically predicted ALS risk are unlikely to be dependent on early or late stage of ALS.

By using summary statistics from several large-scale GWAS, in the present study, we identified a statistically significant positive polygenic association between IgG and ALS in both the discovery and replication analyses. The association is further suggested to be causal in the MR analysis. Although statistically significant result was only obtained in the discovery analysis, similar point estimate was noted in the replication analysis. The differences between the discovery (ALS_2016) and replication (ALS_2018) samples might have contributed to the lack of statistical significance in the replication analysis. For instance, in the discovery GWAS (ALS_2016), patients with ALS and controls were matched for age, sex, and geographic regions, whereas more than 50% of participants in the replication GWAS (ALS_2018) were only matched for race/ethnicity.

In contrast to IgG, no clear polygenic association nor causal relationship was noted for IgA-ALS and IgM-ALS. A recent observational study, including 489 patients with ALS and 17475 neurologically healthy controls, also failed to detect an association between total serum IgA and ALS [20]. The different result pattern between IgA, IgM and IgG might be partly explained by the fact that IgG is a biomarker for chronic inflammation and bypasses the blood-brain-barrier, which might be more relevant for neurodegenerative diseases including ALS, compared to the other immunoglobulins [21]. Besides PRS analyses, we could not perform alternative approaches to test the association between IgG levels and ALS, which could be a potential source for bias. Furthermore, other sensitivity analyses are required to confirm the association between IgG and ALS.

Even though we used the latest and largest GWAS of immunoglobulins, only two independent SNPs could be selected as IVs for IgG. As a result, we could only use IVW methods in the MR analysis. According to the current GWAS Catalog, no genome-wide significant association was identified between the two IVs of IgG and other non-ALS traits. Interestingly, the two genes near these two IVs are both biologically related to immune responses, neuroinflammation, and neurodegeneration. Genetic variant rs7554873 is close to the *FCGR2B* (Fc fragment of IgG receptor IIb) gene on chromosome 1. Protein FcγRIIB encoded by *FCGR2B* gene is a low-affinity receptor for the Fc region of IgG. FcγRIIB involves in the regulation of antibodies produced by B-cells and in the phagocytosis of immune complexes, it is also reported to mediate the inhibitory effect of aggregated α-synuclein on microglial phagocytosis [22]. Genetic variant rs2133037 is close to the HLA-DQB1 (major histocompatibility complex, class II, DQ beta 1) gene on chromosome 6. Its encoded protein HLA-DQB1 is also reported to be associated with immunity and neuronal survival [23].

In this study, we had only access to GWAS summary statistics data when testing the polygenic and causal relationship between serum immunoglobulins (IgA, IgM, IgG) and ALS. Using a Mendelian randomization analysis, our study is less prone to methodological limitations commonly seen in observational studies such as confounding and reverse causation. Future studies with individual genotype data are warranted in further understanding the roles of other factors (such as age, sex, ethnicity, etc.) in the link between different immunoglobulins (especially IgG) and ALS. More importantly, to further confirm the causal association between IgG and ALS, it should remove the possibility that IgG is a biomarker of ALS rather than a risk factor of ALS. The co-localization analysis for each instrumental variable of IgG might also be helpful to identify if the causal association is driven by common causal genetic variants between IgG and ALS.

Further, to test whether chronic inflammation is indeed a mechanism linking together IgG and ALS, the role of IgG on other diseases (both diseases with a known link with chronic inflammation and diseases without such a link) needs to be studied. Finally, as high levels of IgG antibodies were suggested to improve the diagnosis of ALS [19], the potential use of IgG measurement in clinical diagnosis of ALS needs to be further examined.

## Conclusions

In conclusion, our study suggests a shared polygenic risk between serum IgG and ALS. The causality between IgG and ALS needs to be further validated when more instrumental variables are available and when different methods of causal inference are possible to use.

## Data Availability

GWAS summary statistics of ALS were accessed from the Project MinE group and the ALS Variant Server.

http://als.umassmed.edu/

http://databrowser.projectmine.com/

## List of abbreviations

ALS: Amyotrophic lateral sclerosis
Ig: immunoglobulin
CI: confidence interval
GWAS: genome-wide association study
PRS: polygenic risk score
MR: Mendelian randomization
SNP: single nucleotide polymorphism
OR: odds ratio
LD: linkage disequilibrium
IVW: inverse variance weighted
FCGR2B: Fc fragment of IgG receptor IIb
HLA-DQB1: Major histocompatibility complex, class II, DQ beta 1.

## Declarations

## Acknowledgements

The authors would like to thank the Project MinE group and the ALS Variant Server for sharing the summary statistics of GWAS on ALS.

## Authors’ contributions

X.C. and F.F. contributed to the conception and design of the study; X.C., X.S., X.Z., and Y.Z. contributed to acquisition and analyses of data; X.C., X.S., X.Z.,Y.Z., and F.F. contributed to drafting and revising the manuscript.

## Availability of data and materials

GWAS summary statistics of ALS were accessed from the Project MinE group (https://www.projectmine.com/research/download-data) and the ALS Variant Server (http://als.umassmed.edu).

## Ethics approval and consent to participate

Because GWAS summary statistics rather than individual-level data were used in this study, both informed consent and ethical approval were waived according to the ethical review board in Shenzhen Baoan Women’s and Children’s Hospital.

## Consent for publication

Not applicable.

## References

1. Beers DR, Appel SH: Immune dysregulation in amyotrophic lateral sclerosis: mechanisms and emerging therapies. Lancet Neurol 2019, 18:211–220.

2. Yazdani S, Mariosa D, Hammar N, Andersson J, Ingre C, Walldius G, Fang F: Peripheral immune biomarkers and neurodegenerative diseases: A prospective cohort study with 20 years of follow-up. Ann Neurol 2019, 86:913–926.

3. Vandenbroucke JP, Broadbent A, Pearce N: Causality and causal inference in epidemiology: the need for a pluralistic approach. Int J Epidemiol 2016, 45:1776–1786.

4. Chen X, Yazdani S, Piehl F, Magnusson PKE, Fang F: Polygenic link between blood lipids and amyotrophic lateral sclerosis. Neurobiol Aging 2018, 67: e201–e206.

5. Bandres-Ciga S, Noyce AJ, Hemani G, Nicolas A, Calvo A, Mora G, Consortium I, International ALSGC, Tienari PJ, Stone DJ, et al: Shared polygenic risk and causal inferences in amyotrophic lateral sclerosis. Ann Neurol 2019, 85:470–481.

6. Zhan Y, Fang F: Smoking and amyotrophic lateral sclerosis: A mendelian randomization study. Ann Neurol 2019, 85:482–484.

7. Jonsson S, Sveinbjornsson G, de Lapuente Portilla AL, Swaminathan B, Plomp R, Dekkers G, Ajore R, Ali M, Bentlage AEH, Elmer E, et al: Identification of sequence variants influencing immunoglobulin levels. Nat Genet 2017, 49:1182–1191.

8. van Rheenen W, Shatunov A, Dekker AM, McLaughlin RL, Diekstra FP, Pulit SL, van der Spek RA, Vosa U, de Jong S, Robinson MR, et al: Genome-wide association analyses identify new risk variants and the genetic architecture of amyotrophic lateral sclerosis. Nat Genet 2016, 48:1043–1048.

9. Nicolas A, Kenna KP, Renton AE, Ticozzi N, Faghri F, Chia R, Dominov JA, Kenna BJ, Nalls MA, Keagle P, et al: Genome-wide Analyses Identify KIF5A as a Novel ALS Gene. Neuron 2018, 97:1268–1283.

10. Euesden J, Lewis CM, O’Reilly PF: PRSice: Polygenic Risk Score software. Bioinformatics 2015, 31:1466–1468.

11. Hemani G, Zheng J, Elsworth B, Wade KH, Haberland V, Baird D, Laurin C, Burgess S, Bowden J, Langdon R, et al: The MR-Base platform supports systematic causal inference across the human phenome. Elife 2018, 7:e34408.

12. Armon C: Smoking is a cause of amyotrophic lateral sclerosis. High low-density lipoprotein cholesterol levels? Unsure. Ann Neurol 2019, 85:465–469.

13. Molteni M, Rossetti C: Neurodegenerative diseases: The immunological perspective. J Neuroimmunol 2017, 313:109–115.

14. Novellino F, Sacca V, Donato A, Zaffino P, Spadea MF, Vismara M, Arcidiacono B, Malara N, Presta I, Donato G: Innate Immunity: A Common Denominator between Neurodegenerative and Neuropsychiatric Diseases. Int J Mol Sci 2020, 21(3):1115.

15. Sabatino JJ, Jr., Probstel AK, Zamvil SS: B cells in autoimmune and neurodegenerative central nervous system diseases. Nat Rev Neurosci 2019, 20:728–745.

16. Chen K, Magri G, Grasset EK, Cerutti A: Rethinking mucosal antibody responses: IgM, IgG and IgD join IgA. Nat Rev Immunol 2020, 20(7):427–441.

17. Donnenfeld H, Kascsak RJ, Bartfeld H: Deposits of IgG and C3 in the spinal cord and motor cortex of ALS patients. J Neuroimmunol 1984, 6(1):51–57.

18. Duarte F, Binet S, Lacomblez L, Bouche P, Preud’homme JL, Meininger V: Quantitative analysis of monoclonal immunoglobulins in serum of patients with amyotrophic lateral sclerosis. J Neurol Sci 1991, 104(1):88–91.

19. May C, Nordhoff E, Casjens S, Turewicz M, Eisenacher M, Gold R, Brüning T, Pesch B, Stephan C, Woitalla D, et al: Highly immunoreactive IgG antibodies directed against a set of twenty human proteins in the sera of patients with amyotrophic lateral sclerosis identified by protein array. PLoS One 2014, 9(2):e89596.

20. Crayle J, Elmallah M, Sleasman J, Bedlack R: Total serum immunoglobulin A in ALS. Amyotroph Lateral Scler Frontotemporal Degener 2020, 22(1-2):61–65.

21. Watson N, Ding B, Zhu X, Frisina RD: Chronic inflammation - inflammaging - in the ageing cochlea: A novel target for future presbycusis therapy. Ageing Res Rev 2017, 40:142–148.

22. Choi YR, Kang SJ, Kim JM, Lee SJ, Jou I, Joe EH, Park SM: FcgammaRIIB mediates the inhibitory effect of aggregated alpha-synuclein on microglial phagocytosis. Neurobiol Dis 2015, 83:90–99.

23. Hallberg P, Smedje H, Eriksson N, Kohnke H, Daniilidou M, Ohman I, Yue QY, Cavalli M, Wadelius C, Magnusson PKE, et al: Pandemrix-induced narcolepsy is associated with genes related to immunity and neuronal survival. EBioMedicine 2019, 40:595–604.

